# Optimal strategies for adapting open-source large language models for clinical information extraction: a benchmarking study in the context of ulcerative colitis research

**DOI:** 10.1101/2024.11.06.24316817

**Authors:** Richard P. Yim, Anna L. Silverman, Shan Wang, Vivek A. Rudrapatna

## Abstract

**Background:** Closed-source large language models (LLMs) like GPT-4o have shown promise for clinical information extraction but are potentially limited by cost, data security concerns, and inflexibility. Open-source models have emerged as an attractive alternative, with many LLM adaptation strategies developed in the literature. However, it is currently unclear what adaptation strategies are optimal, and how they ultimately compare to closed-source models.

**Methods:** We studied the effects of three common LLM adaptation strategies: chain-of-thought prompting, few-shot prompting, and fine-tuning. Our target for information extraction was the Mayo Endoscopic Subscore (MES). We applied those strategies in all combinations to six open-source models (8-70 billion parameters) using an annotated set of colonoscopy procedure reports from two centers: the University of California, San Francisco (N=608) and San Francisco General Hospital (N=217). We analyzed the relationship of these strategies to several performance metrics with a mixed-effects model, accounting for the variability between centers and LLMs. GPT-4o was not subject to QLoRA due to its closed-source nature but was used as a comparator in our benchmarks. We also provide in-depth commentary on the cost-effectiveness of these open-source LLMs and GPT-4o for MES extraction.

**Results:** Across adaptation strategies, QLoRA statistically (*p* < 0.001) improves the performance of open-source LLMs by 8.3-15.6 percentage points across accuracy, precision and recall. However, GPT-4o with prompt engineering is superior to the best open-source model by a margin of 2.5-5.4%. Yet, a simple cost-effectiveness analysis suggests that GPT-4o is expensive compared to open-source models.

**Conclusion:** GPT-4o is currently the most performant LLM for MES extraction. If unavailable, open-source models optimized with QLoRA are a competitive alternative. However, our results also suggest that current instruction-following LLMs including GPT-4o do not fully follow user-provided instructions, leaving room for improvement. More work is needed to achieve consistent, near-perfect performance in clinical information extraction by LLMs.

**Brief Description:** In this study, we compare interactions of finetuning, chain-of-thought prompting, and few-shot prompting on open-source models to extract useful information from clinical notes. We conclude that QLoRA results in statistically and practically significant performance gains across several metrics while other strategies and their combinations have little impact.

## Introduction

Electronic health records (EHR) data has tremendous potential to be useful for studying and improving the practice of medicine. However, important information in the EHR is found in clinical notes. Robust and cost-effective methods for extracting information from clinical notes are an important step towards realizing the full value of EHR data. Closed-source large language models (LLMs) like GPT-4o perform well on medical knowledge tasks, but lack in ability with respect to information extraction [1, 2]. Open-source LLMs are emerging as potential alternatives to closed-source LLMs. Open source LLMs are attractive due to improved security for protected health information (PHI) data. It is unknown how open-source LLMs should be optimized or adapted for clinical information extraction [3, 4]. Additionally, it is unclear how cost-effective current open-source models are compared to GPT-4o for clinical information extraction.

We investigate these questions in the context of ulcerative colitis (UC), a chronic gastrointestinal disorder. UC is commonly characterized by the Mayo endoscopic subscore (MES), a quantitative score that corresponds directly to the mucosal inflammatory activity of the colon of a patient affected by UC [5]. The MES can be readily extracted from colonoscopy procedure reports (i.e., free-text data). Quantitative scores like the MES are better suited to study real-world treatment effects and improve patient care in this disease [6]. Importantly, we emphasize the distinction between information retrieval and information extraction, as the MES value must be extracted using a complex and nuanced annotation protocol, a task that is well-suited for LLMs. We primarily accomplish two things in this paper, within the context of UC: (1) benchmarking current state-of-the-art (SoA) LLMs,(2) statistically identifying the best LLM adaptation strategies for MES extraction.

## Methods

### Source Data

We used a set of colonoscopy reports from University of California, San Francisco (UCSF; N=608) and San Francisco General Hospital (SFGH; N=217) from Silverman et al [7]. These reports, written between 2017-2020, corresponded to patients with ulcerative colitis diagnosis codes and were annotated by two clinical experts according to the MES, an ordinal scale of UC activity ranging from 0 (quiescent) to 3 (severe). Scores of “-1” were assigned to reports that should not be annotated according to the MES due to a diagnosis of Crohn’s disease or a history of surgical alterations. To improve the class balance prior to downstream evaluations, we annotated an additional 327 reports at UCSF. This resulted in a total of 825 human-annotated notes with at least 100 samples in each of the five classes.

To measure the annotation quality of this augmented corpus, we resampled six notes from each class in each corpus (UCSF and SFGH), totaling 60 notes altogether. These were re-annotated in a blind fashion by both original annotators. We report interrater reliability 1.0 and 0.83 for UCSF and SFGH, respectively, using Cohen’s kappa [8]. Finally, we partitioned the UCSF data according to a 70/30 train-test split (425 training notes and 183 testing notes). Due to data access limitations, we were unable to acquire additional training data from the SFGH corpus. The distributions of our datasets in each class and average lengths of reports in each corpus can be found in Table S1, Supplementary Appendix.

### Large Language Models and Benchmarking

We selected a total of eight LLMs to assess their abilities for clinical information extraction in the presence of model adaptation strategies. We selected one popular closed-source model, GPT-4o-2024-05-13 (GPT-4o), and six open-source, instruction-tuned LLMs: Gemma-2-9b-it, Llama3.1-8b-instruct, Phi-3-medium-128K-instruct, Mistral-Nemo-Instruct-2407, Gemma-2-27b-it, Llama3.1-70b-instruct, and Mistral-Large-Instruct-2407. With the exception of Mistral-Large-Instruct-2407 at a 123 billion parameters, open-source models were selected based on their small-to-medium capacity (8-70 billion parameters) and reflect our judgment of what models can be finetuned and are deployable at well-resourced medical centers. The execution details of these models can be found in the Supplementary Appendix. We accessed GPT-4o via a protected health information (PHI) secure API provisioned by UCSF.

To benchmark models, we calculated the following metrics on a single test set: unweighted accuracy, macro-averaged precision, and macro-averaged recall. Additionally, our analysis includes practical performance measures such as a MES scorable accuracy (i.e., accuracy of “-1” class), MES scorable precision, and under-classification rate. The MES scorable accuracy reflects whether LLMs can correctly abstain from assigning an MES, and the MES scorable precision indicates how often LLMs commit harmful false negative errors. To elaborate, a low MES scorable precision indicates that the LLM more often than not classifies MES scorable notes as being not scorable effectively throwing away information. The under-classification rate communicates how often colonoscopy reports are annotated with dangerous, lower-than-expected MES values. For example, a colonoscopy report corresponding to an MES of 3, the highest level of UC disease activity, should not at all be classified by an LLM with having an MES of 0, or nonexistent UC disease activity.

### Experimental Design for Identifying Optimal LLM Adaptation Strategies

To comprehensively study the impact of model adaption strategies on performance, we used an unreplicated 2^3^ full factorial experimental design with complete blocking by center and model [9]. This design accounts for all possible interactions of three strategies: no finetuning versus quantized low-rank adaptation (QLoRA) finetuning; no CoT prompting versus zero-shot-CoT prompting; and zero-shot versus five-shot prompting. We specifically use QLoRA finetuning to minimize hardware requirements and optimize finetuning efficiency [10, 11]. We use zero-shot-CoT prompting which has shown to be as effective as conventional full one-shot-CoT prompting [12]. We specifically use five-shot prompting as it corresponding to an example from each of the five classes in our data [13]. For five-shot prompting and finetuning, we use the training set of UCSF notes as source data. We have complete blocking since we execute the full factorial design across each center and each LLM. We were unable to finetune GPT-4o and Mistral-Large-Instruct-2407 due to API restrictions and hardware memory limitations, respectively. Therefore, those two models, while benchmarked, are excluded in our statistical model (see the following section). Figure 1A shows the workflow and model development pipeline and Figure 1B illustrates the experimental design. Full technical implementation details can be found in the Supplementary Appendix.

**Figure 1:**
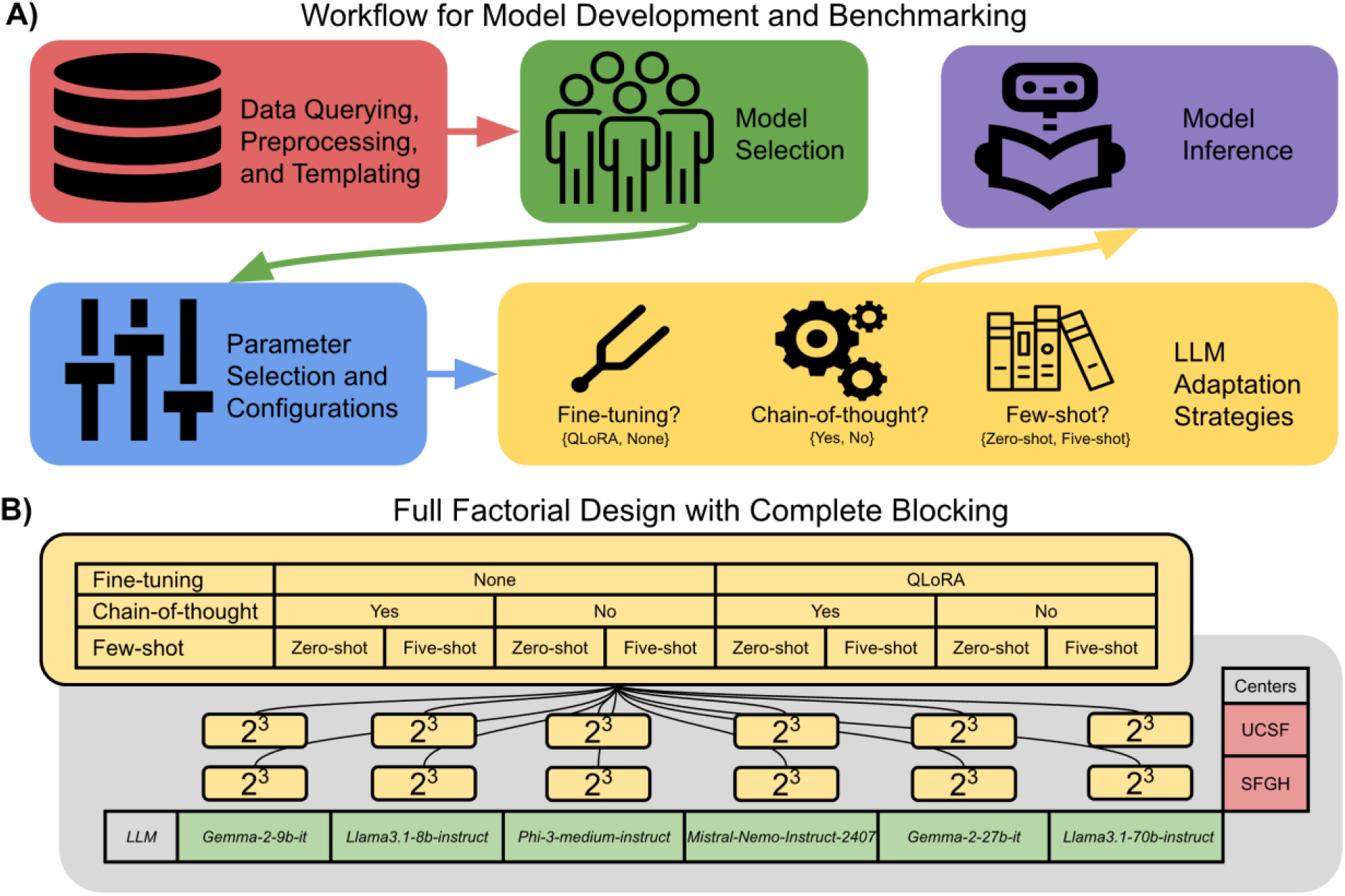
Diagram A illustrates our workflow from data querying to LLM inference. Diagram B illustrates our experimental design where we test and produce inferences for all interactions among our three adaptation strategies at two levels (i.e., 2^3^ = 8 unique binary strings of length 3 in each block). We show blocking by rows corresponding to data from two medical centers and by columns corresponding to inferences made by six different open-source models. Therefore, we have a total of 96 measured experimental units (not including benchmarks for GPT-4o-2024-05-13 and Mistral-Large-2407).

### Mixed-Effects Model of LLM Adaptation Strategies

With our experimental design, we recorded all LLM annotations with all LLM adaptation strategies on both datasets. With the LLM annotations, we calculated and statistically modeled four response variables: unbalanced accuracy, macro-averaged precision, macro-averaged recall, and MES scorable accuracy. Again, we consider our blocks to be random effects and our LLM adaptation strategies to be fixed effects that can be controlled by an investigator. With these four response variables, we generated four mixed-effects models according to the following formulation:

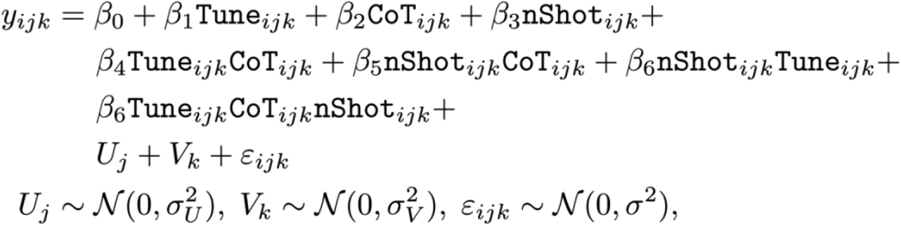

where *j* indexes each center (UCSF, SFGH); *k* indexes each of the six open-source LLMs (Gemma-2-9b-it, Llama3.1-8b-instruct, Phi-3-medium-128K-instruct, Mistral-Nemo-Instruct-2407, Gemma-2-27b-it, Llama3.1-70b-instruct); and *i* indexes each fixed effect interaction within each (*j, k*)-block [14]. The variable names are self-commenting. We model these strategies with two random intercepts corresponding to each block dimension (*U*_*j*_ and *V*_*k*_), and we have a residual term *ϵ*_*ijk*_—these variables are normally distributed with their own variances. Finally, *y*_*ijk*_ corresponds to the *i*^th^ treatment configuration in block (*j, k*), of which there are eight total. Further commentary on model diagnostics and mixed effects modeling choices are provided in the Supplementary Appendix.

## Results

### Benchmarking LLMs and Adaptation Strategies

Model performance metrics are reported in Table 1. The best-performing model on average over all adaptation strategy combinations and over both datasets is GPT-4o13 in accuracy (79.6%), macro-averaged recall (78.0%), and MES scorable accuracy (92.1%). We note that Mistral-Large-Instruct-2407 performed similarly to GPT-4o in macro-averaged precision (78.6% vs 78.1%). These model comparisons, however, are not one-to-one as GPT-4o and Mistral-Large-Instruct-2407 were not finetuned for technical reasons, unlike the smaller open-source models (<100B parameters). Amongst the smaller LLMs, Meta-Llama-3.1-70B-Instruct follows closely behind the larger Mistral-Large-Instruct-2407 model and performs similarly across other metrics (74% average accuracy).

**Table 1:** Average performance of each model across all prompt strategies. For models under 100B parameters all values are averaged with N=16 corresponding to eight LLM adaptation strategy combinations over two datasets, UCSF and SFGH. For the models over 100B parameters, GPT-4o-2024-05-13 and Mistral-large-Instruct-2407, N=8 as these models were not finetuned. We provide standard deviations in parentheses. (The best-performing models on average are asterisked and corresponding metrics are underlined.)

| Large Language Model | Average Accuracy | Average Precision | Average Recall | Average MES scorable Accuracy | Average Total Time in Hours | Average Total Cost in USD |
| --- | --- | --- | --- | --- | --- | --- |
| GPT-4o-2024-05-13 (>100B)* | <u>79.6</u> (6.5) | 78.1 (5.1) | <u>78.0</u> (8.3) | <u>92.1</u> (5.6) | 0.1 (0.1) | 4.4 (2.2) |
| Mistral-Large-Instruct-2407 (123B)* | 74.7 (7.7) | <u>78.6</u> (4.2) | 75.5 (7.2) | 86.7 (8.4) | 1.0 (0.5) | 3.6 (2.0) |
| Meta-Llama-3.1-70B-Instruct (70B) | 74.0 (7.6) | 75.7 (5.1) | 72.9 (6.3) | 88.5 (8.0) | 2.7 (2.2) | 9.8 (8.1) |
| Meta-Llama-3.1-8B-Instruct (8B) | 65.9 (11.8) | 65.7 (10.9) | 62.5 (12.4) | 83.1 (10.7) | 0.6 (0.4) | 0.3 (0.2) |
| Mistral-Nemo-Instruct-2407 (12B) | <u>64.2</u> (11.1) | 67.1 (6.9) | <u>64.2</u> (9.8) | 82.1 (12.3) | 0.8 (0.6) | 0.4 (0.3) |
| Phi-3-medium-128k-instruct (14B) | 59.7 (12.6) | 61.5 (9.5) | 59.2 (12.2) | 79.0 (13.1) | 1.0 (0.7) | 0.5 (0.3) |
| Gemma-2-9b-it (9b) | 54.5 (14.0) | 58.4 (12.0) | 59.7 (11.1) | 76.3 (10.8) | 0.8 (0.6) | 0.4 (0.3) |
| Gemma-2-27b-it (27b) | 52.1 (12.1) | 56.2 (9.7) | 59.5 (9.4) | 71.4 (10.7) | 1.3 (0.9) | 4.7 (3.5) |

Tables 2 shows relevant performance benchmarks of each adaptation strategy combination (QLoRA finetuning, Zero-shot-CoT prompting, five-shot prompting). We found that for models under 100B parameters, QLoRA in combination with zero-shot prompting is the optimal adaptation strategy for MES extraction. There are negligible performance gains between five- and zero-shot prompting. Consequently, we argue that zero-shot prompting is most optimal since it performs comparably to five-shot prompting with fewer token inputs. For LLMs greater than 100B parameters, LLMs that were not finetuned in our experiments, we find that zero-shot prompting without chain-of-thought makes for optimal prompting. Full unabridged performance breakdowns, and all results and measures from our experiments can be found in Table S2 and Table S3, Supplementary Appendix, for UCSF and SFGH, respectively. Our statistical modeling results in the next section can be replicated using data from these tables.

**Table 2:** Average stratified performance for each prompt strategy combination. For large language models with less than 100B parameters, averaging is done over N=12 samples, corresponding to six models and two datasets. For LLMs over 100B parameters averaging is done over N=4 samples, corresponding to two models and two datasets. We provide standard deviations in parentheses. (The best-performing combination of strategies are asterisked and corresponding metrics are underlined.)

| Large-Language with Less than 100B Parameters<br>(Gemma-2-9b-it, Llama3.1-8b-instruct, Phi-3-medium-128K-instruct, Mistral-Nemo-Instruct-2407, Gemma-2-27b-it, Llama3.1-70b-instruct) |  |  |  |  |  |  |  |  |
| --- | --- | --- | --- | --- | --- | --- | --- | --- |
| n-Shot | Tuning | Chain-of-Thought | Average Accuracy | Average Precision | Average Recall | Average MES Scorable Accuracy | Average Total Time | Average Total Cost |
| Five-shot | QLoRA | No | 57.0 (16.2) | 61.0 (14.7) | 58.7 (14.1) | 74.5 (14.1) | 1.9 (1.3) | 4.3 (6.3) |
|  |  | Yes | 61.1 (13.3) | 63.7 (11.5) | 62.2 (11.9) | 79.3 (13.0) | 2.3 (1.4) | 5.2 (7.3) |
|  | None | No | 59.1 (13.0) | 61.7 (11.0) | 62.3 (9.9) | 76.0 (11.3) | 0.2 (0.1) | 0.4 (0.5) |
|  |  | Yes | 57.5 (13.7) | 60.7 (10.7) | 60.2 (9.7) | 76.8 (12.1) | 0.5 (0.2) | 0.9 (1.1) |
| Zero-shot* | QLoRA* | No* | 73.0 (10.1) | <u>73.1 (8.2)</u> | 70.7 (9.3) | <u>90.0 (8.0)</u> | 1.8 (1.3) | 4.1 (6.0) |
|  |  | Yes* | <u>73.2 (10.3)</u> | 72.0 (9.3) | <u>71.3 (10.0)</u> | <u>90.0 (8.4)</u> | 2.2 (1.4) | 4.9 (7.0) |
|  | None | No | 57.3 (10.4) | 62.2 (7.8) | 62.4 (8.3) | 78.8 (8.7) | 0.1 (0.1) | 0.3 (0.3) |
|  |  | Yes | 55.8 (9.0) | 58.4 (6.9) | 56.2 (8.8) | 75.0 (9.7) | 0.6 (0.3) | 1.1 (1.4) |
| Large-Language with Greater than 100B Parameters<br>(Mistral-Nemo-Large-2407, GPT-4o-2024-05-13) |  |  |  |  |  |  |  |  |
| n-Shot | Tuning | Chain-of-Thought | Average Accuracy | Average Precision | Average Recall | Scorable Accuracy | Average Total Time | Average Total Cost |
| Five-shot | None | Yes | 75.5 (7.6) | 76.8 (2.7) | 75.1 (6.1) | 88.9 (9.0) | 0.8 (0.8) | 6.0 (1.2) |
|  |  | No | 79.5 (8.5) | 80.0 (5.2) | 79.3 (8.4) | 90.8 (8.2) | 0.3 (0.3) | 4.1 (2.4) |
| Zero-shot* | None | Yes | 72.0 (5.7) | 75.3 (5.4) | 70.2 (6.2) | 86.3 (9.5) | 0.8 (0.8) | 3.9 (1.7) |
|  |  | No* | <u>81.4 (5.9)</u> | <u>81.2 (3.0)</u> | <u>82.3 (5.8)</u> | <u>91.6 (4.0)</u> | 0.2 (0.2) | 1.9 (0.6) |

### Statistical Findings for Adaptation Strategies

We estimated a 3^rd^ order mixed-effects model corresponding to all interactions between our three adaptation strategies of interest: QLoRA finetuning, Zero-shot-CoT, and five-shot prompting. We modeled these effects against unweighted accuracy, macro-averaged precision, macro-averaged recall, and MES scorable accuracy resulting in four separate mixed-effects models. Figure 2A visualizes the random intercept estimates for each LLM and center. We plot the estimates and 95% confidence intervals of the random intercepts as rectangles. Consistently, we found better MES extraction performance on UCSF data compared to SFGH, which is consistent with our reported interrater reliability measure—we achieved perfect IRR for the UCSF dataset and have better UCSF inferences compared to SFGH, which has high but imperfect IRR. We observe decreasing LLM performance as the number of parameters in billions is increased, until we get to Llama3.1-70B-Instruct with 70B parameters where there is a modest return to increase in performance across our four metrics. We also provide the caveat that these intercept estimates are confounded with the residuals from our mixed-effects model specification, and do not strongly correspond to hard differences in model performance although these trends are reflected in our benchmarks (e.g., Meta-Llama3.1 family of models perform the best among small-medium size models, and MES extraction is easier on UCSF data).

**Figure 2:**
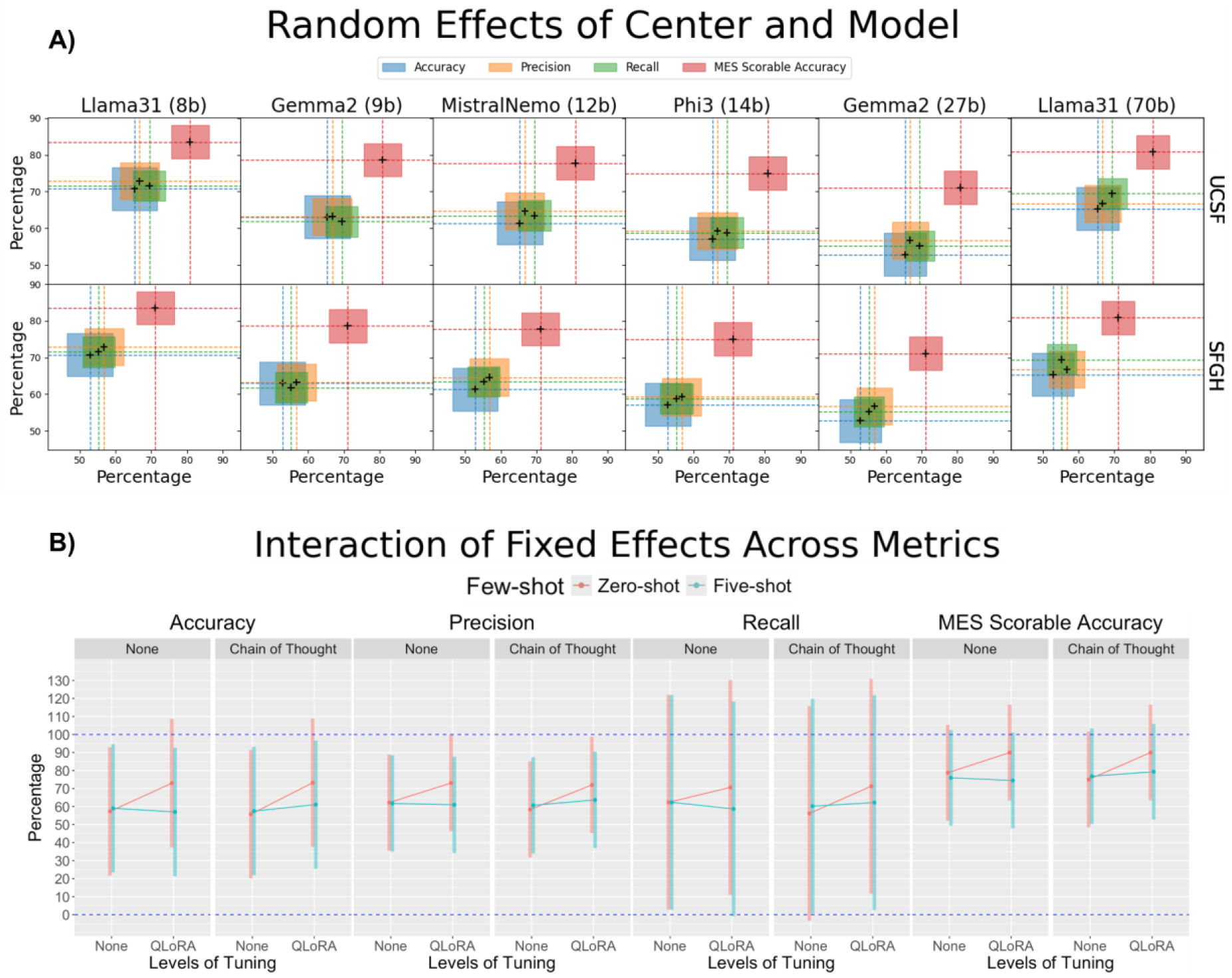
Visualization of fixed effects. Plot A shows the random intercepts adjusted by the estimated baseline fixed effect shown in our linear model formulation across all four response variables: accuracy, macro-averaged precision, macro-averaged recall, and MES scorable accuracy. Plot B shows interactions of estimated marginal means, with corresponding 95% confidence intervals, between few-shot prompting and finetuning stratified by chain-of-thought prompting, again across our four response measurements. Both plots must be interpreted conservatively, although they reflect what we observe and understand from our benchmarks.

From our mixed effect model estimates, we found that the only statistically significant (*α* = 0.05) and consistent adaptation strategy was QLoRA finetuning resulting in an estimated 8.3-15.6 percent *increase* in performance across our four models, holding all other effects constant. The only interaction that was found to be statistically significant between all four mixed-effects models was the interaction between QLoRA finetuning and few-shot prompting resulting in a 11.9-17.7 percent *decrease* across our four performance measures, again, holding all other effects constant. Figure 2B illustrates the estimates of marginal means between few-shot prompting and LLM finetuning stratified by CoT. Due to the single replicate nature of our experiment, these plots must be conservatively interpreted—we elaborate on the very wide confidence intervals in the discussion section as a limitation of our design. We see that with QLoRA finetuning, there is an increase in measured performance across each response type. The only observed exception is with five-shot prompting without Zero-shot-CoT, where we found that there is a consistent marginal decrease in all performance measures with QLoRA finetuning.

We performed additional post hoc analysis to estimate marginal mean differences among statistically significant fixed effects. The model estimates for each statistically significant fixed effect are reported in Table 3, and we also report corresponding statistically significant pairwise marginal mean differences using Tukey’s Honestly Significant Difference (HSD) test [15]. We can see in Table 3 that the variation in the first-order effect of QLoRA tuning is confounded with the second-order interactions being statistically significant. From our pairwise test, we see that the difference in marginal means between QLoRA and no QLoRA ranges from 5.4-8.7 points across our four performance measures; however, we see additional significant variation from the pairwise differences in the interaction between QLoRA and few-shot prompting. Although statistically significant, as illustrated by very wide confidence intervals in Figure 2B these interpretations must be taken with appropriate caution.

**Table 3:** We show statistically significant estimates of fixed effects as well as statistically significant pairwise differences in estimated marginal means across those fixed effects. Again, note that for our recall response model, we find Zero-shot-CoT and QLoRA with Zero-shot-CoT to be statistically significant. All estimates appear to practically significant as well with effect sizes generally being above five points across all effects and pairwise differences. Again, pairwise difference from estimated marginal means must be conservatively accepted due to the single replicate nature of our experimental design.

| Response | Value | Variables and Pairwise Differences | Estimate (p-value) |
| --- | --- | --- | --- |
| Accuracy | Mixed-effect model estimates | (Intercept) | 57.3 ($p \approx 2.2e-02$) |
| | | QLoRA | 15.6 ($p \approx 6.1e-07$) |
| | | QLoRA:Five-shot | -17.7 ($p \approx 4.1e-05$) |
| | Pairwise differences from estimated marginal means | (QLoRA) – (No QLoRA) | 8.7 ($p < 0.001$) |
| | | (QLoRA, Zero-shot) – (No QLoRA, Zero-shot) | 16.6 ($p < 0.001$) |
| | | (QLoRA, Zero-shot) – (No QLoRA, Five-shot) | 14.8 ($p < 0.001$) |
| | | (QLoRA, Zero-shot) – (QLoRA, Five-shot) | 14.1 ($p < 0.001$) |
| MES Scorable Accuracy | Mixed-effect model estimates | (Intercept) | 78.8 ($p \approx 6.6e-03$) |
| | | QLoRA | 11.2 ($p \approx 8.9e-04$) |
| | | QLoRA:Five-shot | -12.7 ($p \approx 7.1e-03$) |
| | Pairwise differences from estimated marginal means | (QLoRA) – (No QLoRA) | 6.8 ($p < 0.001$) |
| | | (QLoRA, Zero-shot) – (No QLoRA, Zero-shot) | 13.1 ($p < 0.001$) |
| | | (QLoRA, Zero-shot) – (No QLoRA, Five-shot) | 13.6 ($p < 0.001$) |
| | | (QLoRA, Zero-shot) – (QLoRA, Five-shot) | 13.1 ($p < 0.001$) |

### Clinical Information Extraction Cost-Effectiveness

Our whole experiment was very costly. We recorded the average time and cost for LLM inference and finetuning in Tables 1 and 2. Summing all dollar costs required to run a single replicate as we have done across various compute resources, we spent $318.70. To replicate each interaction across several folds of train-test splits is a nontrivial monetary cost. Further, summing all compute hours dedicated to producing all results including finetuning open-source LLMs, we spent 124.12 hours for a single replicate. It is easy to suggest running several replicates in our design, and with these unignorable costs, the decision to precisely make out an additional number of desired replicates may be a very pathological calculation. The full unabridged expense columns for our entire experiment can be found in Tables S2 and S3, in the Supplemental Appendix; cost calculations in full detail can also be found in the Supplemental Appendix.

Meta-Llama-3.1-8B-Instruct was the most efficient model on average across all tests of smaller models (<20B parameters) that had been finetuned, according to Table 1. More precisely, we find that there is an average accuracy to average dollar ratio of 219.6 compared to 18.1 for GPT-4o. While this degree of cost efficiency may appear impressive, this difference may be less important as high quality clinical information extraction performance is worth every increase in monetary expenses. Further, the absolute dollar amount for annotation on a total of 400 notes from UCSF (N-test=183) and SFGH (N-test=217) combined totals less than $4.40 on average using GPT-4o. Scaling this amount to 100 times as many notes, or 40,000 notes, this would cost roughly $450 to make API calls to GPT-4o versus $30 with Meta-Llama-3.1-8B-Instruct using a cloud compute solution.

Anecdotally, we report experiencing substantial technical and implementation issues compounded by hardware limitations. Our development hours and compute costs for development are difficult to quantify, yet very real. Many of the previously cited studies do not use real-world PHI data for clinical information extraction as we do. For example, our access to OpenAI LLMs was provisioned by UCSF IT, and we used a private and secure local machine for our open-source experiments, and not some allegedly private cloud compute service like Google Colab. Other researchers interested in utilizing other closed-source models must be mindful of this detail. Therefore, the real human resource cost—from the proper IT and network security teams to medically trained annotators—, and hardware resource expenses at a medical center to deploy GPT-4o are substantial hidden costs not formally included in our calculations.

## Discussion

### Limitations and Future Work

From our mixed-effects model analysis we found that the interaction of five-shot prompting and QLoRA decreases MES extraction quality. A simple explanation is that since we finetuned our data on zero-shot completions, and the longer five-shot prompts induce bias in the finetuned LLM at inference time. Of course, one can easily suggest finetuning a model with few-shot prompts, but again there are important cost and time considerations with finetuning LLMs using five-shot prompting, and it may not at all be feasible with respect to hardware limitations (i.e., memory requirements for longer context windows grows superlinearly).

With such hardware and resource limitations, we also comment on a potential concern from readers. Namely, our experiment is a single replicate design. We only evaluate each interaction in each block exactly once and control LLM inference variation with greedy token sampling. As we have commented earlier, running additional replicates can be done using several train-test splits, but this can be very expensive. Naturally, in our unreplicated design, we observed very wide and illogical confidence intervals in post hoc estimates for all marginal means, and consequently pairwise marginal mean differences. For example, Figure 2B visualizes these marginal means across different levels of interaction but are conservatively interpreted due to very wide confidence intervals—although they were found to be statistically significant from multiple comparisons test. Yet, the substance of our mixed-effects model estimates still remains valid: QLoRA is the most statistically and practically significant LLM adaptation strategy to improve clinical information extraction quality.

Further, there are more statistical modeling analyses that can be conducted from nontrivial experimental designs based on studies from the current literature. From evaluation of LLMs blocked by prompt types to LLM evaluation on exams blocked by subject category there are several studies in the literature that can and should leverage formal statistical modeling to generate rigorous conclusions [16, 17, 18]. Our findings in LLM adaptation strategies are also subject to change over time, yet the statistical tools and methods available to evaluate and discover future best strategies will remain the same.

We additionally note that we were somewhat limited by our datasets. While the target of our study was real-world MES extraction, we could have also annotated data from more centers and additional information extraction targets. There is real-world variability in clinical text between centers. We see this fact reflected from our inter-rater reliability scores as calculated between UCSF and SFGH, and the consequent contrast in performance between centers as was seen in Figure 2A. We hypothesize that more data for finetuning will naturally increase performance in a substantial way, yet we find impressive performance gains from finetuning on a relatively small corpus (425 training notes from UCSF).

## Conclusion

We have designed and evaluated an experiment benchmarking a variety of LLM adaptation strategies with special attention to SoA open-source models for the task of MES extraction on colonoscopy procedure reports from two centers. To the best of our knowledge, while there are several studies benchmarking several adaptation strategies, this is the first study of its kind that goes beyond benchmarking by including formal statistical modeling and analysis. There are countless studies that also benchmark many LLMs in different applications, and the SoA is evolving so much that we argue special attention should be given to techniques and methods for adapting LLMs.

Currently, we cannot recommend using open-source models for MES extraction, and likely clinical information extraction in general. Closed-source models like GPT-4o hosted on closed and secure private networks appear to be more performant. Along the lines of model performance, we find that GPT-4o is the best choice among our selected models for MES extraction even with higher dollar and time costs. We readily speculate from our results that given PHI-compliant accessibility to finetuning platforms for closed-source models, that QLoRA finetuning on models like GPT-4o will demonstrate clinically acceptable (less than a 5% margin of error according to several metrics) for MES extraction and more. Provided a future where researchers have secure access to even more powerful foundation models, we suspect that our statistical and practical findings will remain consistent.

## Data Availability

All data produced in the present work are contained in the Supplemental Appendix. Data used to generate results, patient clinical data, will not be available to others.

## Data Acknowledgement

The authors thank UCSF Academic Research Services for technical support related to enabling software in a secure, PHI compliant environment; UCSF AI Tiger Team for facilitating and managing access to Versa API (UCSF secure access to Microsoft Azure, OpenAI Large Language Models); and the Chancellor’s Task Force for Generative AI.

