## Supplemental Appendix for "Optimal strategies for adapting open-source large language models for clinical information extraction: a benchmarking study in the context of ulcerative colitis research"

### Supplementary Appendix

#### *Large Language Model Inference Parameters*

For model inference we used 4-bit quantization with 'nf4' quant type as well as double quantization. For the datatype we also used brainfloat16. To induce a deterministic behavior in our models for token inference, we set 'num_beams' equal to 1, reducing the next sampled token space to just one token, and set the temperature to 0. This behavior was permitted on all our open-source models from the Hugging Face library allowing for greedy token sampling. For the GPT-4o model from OpenAI, these options are not available. The closest alternative to reduce LLM prompt completion variability was to set temperature to 0, noting that this setting produces a nonzero probability of sampling several likely tokens (i.e., greedy as possible token sampling). To increase inference speed and efficiency we used FlashAttention-2.

#### *Chain-of-thought and Few-shot Templating*

While there are several variations of chain-of-thought prompting, we focused on utilizing zero-shot-chain-of-thought (zero-shot-CoT). This prompting techniques involves prompting the model to both "think step-by-step" as well as explicitly fill complete tokens corresponding to the reasoning behind its prompt completion. We invoked this "think step-by-step" command in all prompts with the main distinction being from the clinical addendum where we explicitly ask the model to provide chain-of-thought reasoning. For few-shot prompting, we added the annotation protocol at the top of the conversation where we provided the annotation protocol as a system prompt if supported by the LLM. Otherwise, we provide the annotation protocol in the first message to the LLM. Then, we alternate between the clinical note and intended text completion.

For modeling zero-shot-CoT in our few-shot prompts, we had an annotator do two things: 1) identify characteristics of the colonoscopy procedure report corresponding to the MES score, and 2) identify the section in the note that this appears. These two fields of information were then combined into a generic tense respecting text template emulating chain-of-thought reasoning. Finally, we used only the zero-shot, completion-only templates in our finetune models. We did this because of GPU memory limitations—five-shot prompting creates very long token sequences—, and to reduce pathological complexity. We finetune our LLMs in one simple and standard way (zero-shot without CoT), and at inference time add zero-shot-CoT and/or five-shot prompting. Table S5 shows our specific conversation templates used across all models.

#### *Large Language Model Finetuning Parameters*

Our training dataset of UCSF colonoscopy procedure reports consisted of 425 notes. Consequently, we had limited clinical notes available for training. Due to the inherent computational complexity of finetuning LLMs over millions of additional parameters, and lack of thousands of data points, we selected a LoRA alpha of adapted to a 90-minute estimated finetuning time, and the conventional rank value of half of the LoRA alpha parameter. We set the alpha to 32 and the rank to 16. For other parameter choices: we use a default LoRA dropout of 0.1; we have targeted all linear modules (q_proj, k_proj, o_proj, v_proj, gate_proj, up_proj, down_proj, lm_head).

For our training process, we performed two-steps of gradient accumulation with batch-size 1, corresponding to an effective batch-size of 2. We use heuristically selected a learning-rate of 2.0e-05 by observing loss trends after a single training batch was completed across several models; we also use a cosine learning rate scheduler. Given the small training set we simply chose 3 training epochs. All code and relevant scripts for reproducibility are publicly available at https://github.com/richpaulyim/Open-Source-LLMs-MES.

##### *Large Language Cost Calculation*

For models with less than 20B parameters, fine-tuning and inference were done on a locally available private and secure machine using an RTX 4090 GPU, so we reported similar costs for utilizing a Tesla T4 using cloud computing on Google Colab (0.7781647/hour). For models with greater than 20B parameters we report costs based on A100 rates with Microsoft Azure virtual machine ($3.67/hour). We reported inference costs for GPT-4o with rates as of September 2024 ($5/million token inputs), which is a function of token length inputs and outputs as opposed to computing time as calculated for our open-source models. The time for inference and LLM finetuning as reported in our manuscript, corresponds to the compute time of the GPU that was utilized. Again, GPT-4o was not finetuned, so the compute time for GPT-4o corresponds instead to the speed of API calls.

##### *Mixed-Effects Model Building*

Since we wanted to understand the effects of common LLM adaptation strategies we built a straightforward third-order linear model. All the LLMs that we used in our study had sufficiently long 128K token context windows, and we suspected that there was essentially no interaction with what we identified as fixed effects in our experiment (e.g., five-shot and zero-shot-CoT prompting). We did try to fit random slopes to our model incorporating an potential confounding by LLM with each of the strategies, but were unable to achieve convergence with restricted maximum likelihood estimation in our mixed-effects model. This, however, is a desirable result since each of these LLMs are trained on massive and diverse text datasets. There is an implicit assumption that no one model would be biased towards any adaptation strategy amongst the ones we experimented for such that there would be a requirement for random slopes. In the same vein, we don't bother characterizing our choice of models as additional regression data, such as model volume, since our random slopes with account for between model variation in totality.

With respect to datasets, again, no one model is known a priori to perform better on one dataset over another. We again resorted to incorporating the effect from our datasets as a random intercept. Additionally, there would be no interaction between any of our adaptation strategies and choice of dataset. These are factors that would naturally have no dependence on each other (between the adaptation strategy and dataset), but since there is variation in our measured inter-rater reliability values, we incorporate a random intercept as additional variation between datasets in model for classification performance. Finally, we report that for $\alpha = 0.05$, using a likelihood ratio test between a model with only one random intercept versus a model with two random intercepts, we found statistically significant nonzero variation in adding an additional intercept term.

##### *Mixed-Effects Model Diagnostics*

With our specified model, we fitted our fixed and random effects against four performance metrics that we measured in our LLM information extraction experiments. After fitting our models, we assessed goodness-of-fit of our linear models. Figure S1 indicates that we have heuristically good linear fit. To elaborate, for each of our four mixed-effects models, we plotted Pearson residuals corresponding to the fitted values of each model and then created a smoothing spline corresponding to a potential nonlinear signal that may indicate poor model fit, and consequently poor model specification. From all four models, seeing that we have relatively flat, and zero-valued splines that our model is heuristically, sufficiently, well specified.

Additionally, we analyzed if there were any outlier issues in our model in two ways. First, we assessed dfbeta values corresponding to the sensitivity of model parameter estimates according to any single data point [1]. Namely, we visualize perturbations in model estimates as a particular data point was left out. Additionally, we plotted Cook's distance values to assess the sensitivity of fitted-values in our models if a data point was left out—again, another heuristic for characterizing outlier influence [2]. For both diagnostics, we observe that there are no consistent data points with high influence across all four-models and both dfbeta plots and Cook's distance plots. Picking standard thresholds for both plots, $2/\sqrt{n}$ and $4/\sqrt{n}$ for dfbeta and Cook's distances plots, respectively, we see that no singular data point is consistently identified as an outlier point across all models. This assessment highlights two features of our experiment: (1) there are likely no practical experimental errors and (2) our models are sufficiently useful in understanding the effects of different LLM adaptation strategies.

#### Supplementary Tables and Figures

**Figure S1:** Goodness-of-fit plots for our mixed effects model across each performance metric as a response. We see among all four mixed-effects models that we have relatively good linear fit with a smoothing spline that appears to show no real nonzero variation in Pearson residuals as a function of fitted values. For the smoothing parameter we choose $\lambda = 1.25$. Each model visually possess sufficient linear fit.

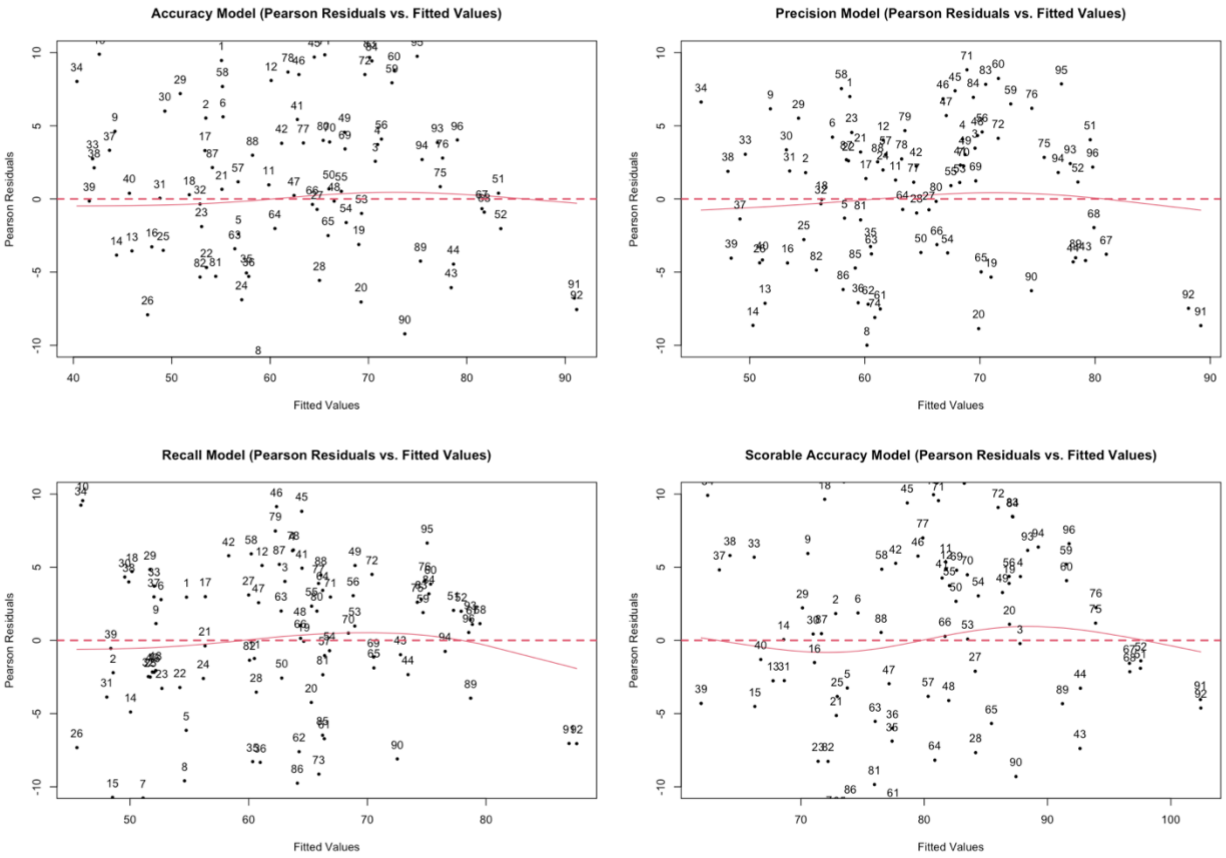

**Figure S2:** We show dfbeta plots of each mixed-effects model for each interaction in each block on the left column and corresponding Cook's distance plots on the right. As shown by the data point indices, we see that there is no singular combination of interactions in every model that is consistently high leverage and high influence. For example, data point 73 in the accuracy row appears to have a high magnitude dfbeta value affecting model estimates, but it does not have appear to appear to have high Cook's distance affecting average fitted values.

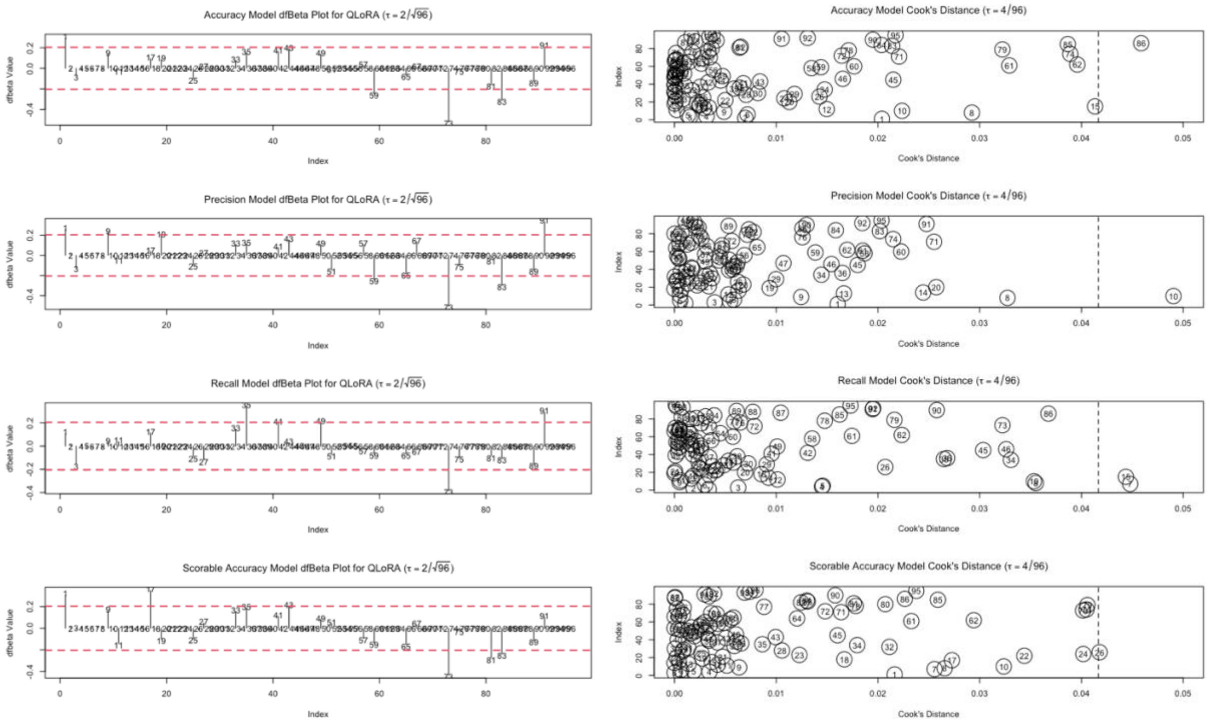

**Table S1:** Centers from two datasets are used in our analysis. Test datasets for UCSF and SFGH as well as one "training" dataset used for both QLoRA finetuning and five-shot prompting. We include average string lengths and across each dataset and string length standard deviation.

| Dataset | Samples | Unscorable (N) | Mayo 0 (N) | Mayo 1 (N) | Mayo 2 (N) | Mayo 3 (N) | Average Length | (SD) |
|---|---|---|---|---|---|---|---|---|
| UCSF Test Set | 183 | 75 (40.98%) | 28 (15.3%) | 26 (14.21%) | 39 (21.31%) | 15 (8.2%) | 2558.91 | 479.85 |
| UCSF Train Set | 425 | 158 (37.18%) | 83 (19.53%) | 52 (12.24%) | 70 (16.47%) | 62 (14.59%) | 2509.0 | 436.56 |
| SFGH Test Set | 217 | 66 (30.41%) | 67 (30.88%) | 20 (9.22%) | 39 (17.97%) | 25 (11.52%) | 8872.84 | 1857.52 |

**Table S2:** Results table from University of California, San Francisco (UCSF) colonoscopy procedure reports for Mayo endoscopic subscore (N=183).

| Model | n-Shot | Tuning | Chain-of-Thought | Accuracy | Precision | Recall | Scorable Accuracy | Scorable Precision | Underclassification | Total Time (hrs) | Total Cost ($) |
|---|---|---|---|---|---|---|---|---|---|---|---|
| Meta-Llama-3.1-8B-Instruct | Zero-shot | None | No | 72.13% | 71.74% | 74.08% | 89.62% | 98.28% | 2.8% | 0.05 hrs | $0.02 |
| | | | Yes | 66.67% | 61.21% | 60.21% | 85.25% | 77.27% | 3.41% | 0.3 hrs | $0.13 |
| | | QLoRA | No | 83.61% | 83.62% | 79.31% | 95.63% | 93.51% | 1.94% | 0.77 hrs | $0.34 |
| | | | Yes | 81.42% | 79.66% | 79.89% | 96.17% | 98.57% | 3.74% | 1.04 hrs | $0.46 |
| | Five-shot | None | No | 68.31% | 69.37% | 69.92% | 83.61% | 89.47% | 1.96% | 0.07 hrs | $0.03 |
| | | | Yes | 66.12% | 63.51% | 66.11% | 87.43% | 94.83% | 2.86% | 0.26 hrs | $0.11 |
| | | QLoRA | No | 67.76% | 68.39% | 67.64% | 85.79% | 86.57% | 1.01% | 0.8 hrs | $0.35 |
| | | | Yes | 75.41% | 74.77% | 71.86% | 91.26% | 87.34% | 2.04% | 1.11 hrs | $0.49 |
| Gemma-2-9b-it | Zero-shot | None | No | 57.92% | 64.84% | 65.36% | 76.5% | 100.0% | 1.85% | 0.08 hrs | $0.04 |
| | | | Yes | 62.84% | 65.51% | 66.13% | 81.42% | 97.67% | 2.8% | 0.43 hrs | $0.19 |
| | | QLoRA | No | 80.33% | 79.17% | 76.59% | 96.72% | 97.26% | 0.94% | 1.02 hrs | $0.45 |
| | | | Yes | 81.42% | 79.84% | 79.18% | 95.63% | 97.18% | 0.94% | 1.39 hrs | $0.61 |
| | Five-shot | None | No | 46.45% | 53.83% | 59.65% | 66.12% | 93.33% | 1.87% | 0.16 hrs | $0.07 |
| | | | Yes | 43.72% | 53.1% | 56.64% | 65.57% | 100.0% | 4.63% | 0.45 hrs | $0.20 |
| | | QLoRA | No | 53.01% | 56.82% | 64.74% | 70.49% | 95.65% | 1.87% | 1.1 hrs | $0.48 |
| | | | Yes | 58.47% | 62.57% | 69.65% | 72.68% | 96.3% | 1.87% | 1.76 hrs | $0.77 |
| Mistral-Nemo-Instruct-2407 | Zero-shot | None | No | 63.39% | 65.12% | 68.67% | 79.78% | 89.58% | 1.94% | 0.05 hrs | $0.02 |
| | | | Yes | 63.93% | 63.14% | 64.51% | 81.97% | 83.87% | 2.04% | 0.37 hrs | $0.16 |
| | | QLoRA | No | 80.87% | 77.19% | 79.94% | 95.08% | 97.14% | 1.89% | 1.18 hrs | $0.52 |
| | | | Yes | 80.87% | 77.94% | 80.63% | 94.54% | 98.51% | 1.87% | 1.55 hrs | $0.68 |
| | Five-shot | None | No | 71.04% | 70.87% | 69.39% | 87.43% | 85.14% | 2.06% | 0.37 hrs | $0.16 |
| | | | Yes | 69.95% | 70.84% | 68.9% | 87.98% | 88.41% | 2.0% | 0.36 hrs | $0.16 |
| | | QLoRA | No | 75.41% | 77.7% | 69.85% | 90.71% | 82.95% | 2.15% | 1.21 hrs | $0.53 |
| | | | Yes | 78.14% | 75.73% | 74.9% | 95.08% | 93.42% | 1.94% | 1.61 hrs | $0.71 |

| | | | | | | | | | | | |
|---|---|---|---|---|---|---|---|---|---|---|---|
| Phi-3-medium-128k-instruct | Zero-shot | None | No | 45.9% | 51.94% | 56.77% | 67.76% | 78.57% | 1.96% | 0.08 hrs | $0.04 |
| | | | Yes | 46.99% | 52.77% | 45.96% | 63.93% | 55.56% | 4.17% | 1.0 hrs | $0.44 |
| | | QLoRA | No | 78.14% | 78.42% | 76.81% | 95.08% | 97.14% | 1.89% | 1.39 hrs | $0.61 |
| | | | Yes | 80.33% | 80.7% | 78.93% | 96.17% | 97.22% | 0.94% | 1.71 hrs | $0.75 |
| | Five-shot | None | No | 67.21% | 65.39% | 69.78% | 86.89% | 96.36% | 1.89% | 0.12 hrs | $0.05 |
| | | | Yes | 70.49% | 65.93% | 69.94% | 90.71% | 98.33% | 1.87% | 0.44 hrs | $0.20 |
| | | QLoRA | No | 73.22% | 68.15% | 69.72% | 93.44% | 90.91% | 1.98% | 1.43 hrs | $0.63 |
| | | | Yes | 69.4% | 66.03% | 67.75% | 93.99% | 98.48% | 2.8% | 1.79 hrs | $0.79 |
| Gemma-2-27b-it | Zero-shot | None | No | 49.18% | 58.2% | 63.9% | 66.12% | 100.0% | 1.85% | 0.16 hrs | $0.59 |
| | | | Yes | 47.54% | 50.93% | 58.71% | 63.93% | 68.0% | 2.0% | 0.5 hrs | $1.84 |
| | | QLoRA | No | 79.78% | 78.33% | 77.35% | 95.63% | 94.67% | 1.92% | 1.83 hrs | $6.72 |
| | | | Yes | 79.78% | 76.37% | 78.36% | 95.63% | 95.89% | 1.9% | 2.22 hrs | $8.14 |
| | Five-shot | None | No | 43.17% | 54.45% | 59.74% | 61.2% | 100.0% | 1.85% | 0.22 hrs | $0.80 |
| | | | Yes | 40.44% | 51.93% | 54.34% | 62.84% | 100.0% | 4.63% | 0.51 hrs | $1.89 |
| | | QLoRA | No | 56.28% | 61.08% | 67.78% | 72.13% | 100.0% | 1.85% | 1.95 hrs | $7.17 |
| | | | Yes | 61.2% | 63.65% | 70.55% | 77.05% | 100.0% | 1.85% | 2.41 hrs | $8.83 |
| Meta-Llama-3.1-70B-Instruct | Zero-shot | None | No | 71.04% | 74.29% | 74.75% | 86.89% | 100.0% | 1.85% | 0.16 hrs | $0.59 |
| | | | Yes | 64.48% | 68.2% | 64.42% | 78.14% | 74.65% | 2.22% | 0.91 hrs | $3.33 |
| | | QLoRA | No | 84.15% | 80.52% | 79.95% | 98.36% | 97.37% | 1.89% | 4.32 hrs | $15.86 |
| | | | Yes | 83.61% | 80.63% | 80.58% | 97.81% | 98.63% | 1.87% | 4.94 hrs | $18.14 |
| | Five-shot | None | No | 80.87% | 80.26% | 80.08% | 94.54% | 97.1% | 1.89% | 0.25 hrs | $0.92 |
| | | | Yes | 78.14% | 78.6% | 75.8% | 95.63% | 97.18% | 2.83% | 0.69 hrs | $2.52 |
| | | QLoRA | No | 84.7% | 84.94% | 81.69% | 98.36% | 97.37% | 1.89% | 4.47 hrs | $16.41 |
| | | | Yes | 83.06% | 81.97% | 79.08% | 98.36% | 96.15% | 1.9% | 5.07 hrs | $18.60 |
| Mistral-Large-Instruct-2407 | Zero-shot | None | No | 80.87% | 81.41% | 83.34% | 93.44% | 97.01% | 0.0% | 0.34 hrs | $1.26 |
| | | | Yes | 77.6% | 80.43% | 76.15% | 90.16% | 86.08% | 0.0% | 1.22 hrs | $4.49 |
| | Five-shot | None | No | 85.79% | 86.24% | 86.12% | 96.72% | 97.26% | 0.94% | 0.49 hrs | $1.79 |
| | | | Yes | 80.33% | 79.4% | 79.69% | 94.54% | 94.52% | 0.96% | 1.29 hrs | $4.72 |
| GPT-4o-2024-05-13 | Zero-shot | None | No | 87.98% | 84.79% | 89.21% | 95.63% | 98.55% | 0.0% | 0.03 hrs | $1.79 |
| | | | Yes | 69.95% | 67.81% | 63.13% | 95.63% | 95.89% | 16.19% | 0.1 hrs | $2.00 |
| | Five-shot | None | No | 85.79% | 82.15% | 86.11% | 97.27% | 98.61% | 0.0% | 0.06 hrs | $5.25 |
| | | | Yes | 83.06% | 78.73% | 80.71% | 98.36% | 97.37% | 0.94% | 0.1 hrs | $5.79 |

**Table S3:** Results table from San Francisco General Hospital (SFGH) colonoscopy procedure reports for Mayo endoscopic subscore (N=217).

| Model | n-Shot | Tuning | Chain-of-Thought | Accuracy | Precision | Recall | Scorable Accuracy | Scorable Precision | Underclassification | Total Time (hrs) | Total Cost ($) |
|---|---|---|---|---|---|---|---|---|---|---|---|
| Meta-Llama-3.1-8B-Instruct | Zero-shot | None | No | 64.52% | 65.7% | 57.71% | 87.56% | 91.49% | 10.88% | 0.06 hrs | $0.03 |
| | | | Yes | 58.99% | 56.66% | 46.37% | 74.65% | 54.95% | 11.88% | 0.38 hrs | $0.17 |
| | | QLoRA | No | 73.27% | 73.04% | 67.08% | 87.56% | 72.94% | 3.91% | 0.81 hrs | $0.36 |
| | | | Yes | 74.65% | 72.58% | 69.82% | 92.17% | 84.51% | 3.57% | 1.12 hrs | $0.49 |
| | Five-shot | None | No | 54.38% | 56.93% | 48.59% | 70.51% | 51.04% | 6.73% | 0.09 hrs | $0.04 |
| | | | Yes | 60.83% | 61.41% | 55.4% | 76.5% | 58.43% | 5.26% | 0.31 hrs | $0.14 |
| | | QLoRA | No | 39.17% | 41.8% | 40.32% | 60.37% | 33.33% | 5.41% | 0.83 hrs | $0.36 |

| | | | | | | | | | | | |
|---|---|---|---|---|---|---|---|---|---|---|---|
| | | | Yes | 47.47% | 50.19% | 45.0% | 64.98% | 44.79% | 4.08% | 1.18 hrs | $0.52 |
| Gemma-2-9b-it | Zero-shot | None | No | 48.85% | 57.96% | 53.34% | 76.5% | 69.23% | 1.44% | 0.11 hrs | $0.05 |
| | | | Yes | 52.53% | 60.17% | 55.56% | 80.18% | 72.55% | 2.92% | 0.59 hrs | $0.26 |
| | | QLoRA | No | 60.83% | 63.96% | 59.25% | 87.1% | 75.0% | 2.27% | 1.06 hrs | $0.47 |
| | | | Yes | 68.2% | 65.62% | 66.24% | 86.64% | 74.03% | 1.53% | 1.54 hrs | $0.68 |
| | Five-shot | None | No | 42.4% | 44.21% | 50.11% | 64.98% | 35.29% | 1.55% | 0.21 hrs | $0.09 |
| | | | Yes | 40.55% | 41.65% | 45.15% | 68.66% | 46.15% | 4.38% | 0.59 hrs | $0.26 |
| | | QLoRA | No | 30.41% | 36.94% | 37.8% | 61.75% | 32.65% | 1.69% | 1.16 hrs | $0.51 |
| | | | Yes | 44.7% | 48.91% | 49.85% | 69.59% | 50.0% | 1.61% | 1.67 hrs | $0.74 |
| Mistral-Nemo-Instruct-2407 | Zero-shot | None | No | 56.68% | 61.49% | 59.33% | 88.02% | 80.3% | 1.45% | 0.07 hrs | $0.03 |
| | | | Yes | 52.07% | 56.22% | 54.87% | 81.57% | 66.25% | 1.61% | 0.44 hrs | $0.19 |
| | | QLoRA | No | 65.9% | 65.62% | 64.58% | 90.78% | 84.85% | 2.13% | 1.21 hrs | $0.53 |
| | | | Yes | 62.21% | 61.03% | 61.04% | 88.02% | 87.04% | 1.39% | 1.62 hrs | $0.71 |
| | Five-shot | None | No | 55.76% | 62.84% | 55.95% | 67.74% | 48.11% | 2.08% | 0.13 hrs | $0.06 |
| | | | Yes | 48.85% | 61.19% | 50.98% | 59.91% | 41.73% | 2.6% | 0.43 hrs | $0.19 |
| | | QLoRA | No | 51.15% | 63.41% | 49.4% | 63.13% | 44.85% | 2.63% | 1.25 hrs | $0.55 |
| | | | Yes | 50.23% | 63.56% | 53.58% | 61.29% | 43.08% | 2.6% | 1.69 hrs | $0.74 |
| Phi-3-medium-128k-instruct | Zero-shot | None | No | 45.62% | 51.93% | 49.2% | 69.12% | 49.32% | 4.39% | 0.11 hrs | $0.05 |
| | | | Yes | 39.63% | 46.5% | 38.2% | 53.92% | 38.19% | 9.68% | 1.0 hrs | $0.44 |
| | | QLoRA | No | 64.06% | 64.83% | 63.1% | 82.03% | 66.27% | 2.44% | 1.43 hrs | $0.63 |
| | | | Yes | 59.45% | 63.54% | 57.1% | 76.5% | 57.01% | 8.57% | 1.82 hrs | $0.80 |
| | Five-shot | None | No | 58.06% | 59.76% | 56.52% | 72.35% | 53.12% | 4.72% | 0.16 hrs | $0.07 |
| | | | Yes | 55.3% | 56.55% | 53.89% | 71.43% | 52.17% | 4.67% | 0.53 hrs | $0.23 |
| | | QLoRA | No | 48.85% | 55.39% | 44.17% | 65.9% | 46.97% | 3.7% | 1.47 hrs | $0.65 |
| | | | Yes | 52.53% | 55.86% | 49.07% | 84.33% | 72.22% | 3.82% | 1.91 hrs | $0.84 |
| Gemma-2-27b-it | Zero-shot | None | No | 44.7% | 52.68% | 55.75% | 71.89% | 64.71% | 1.38% | 0.22 hrs | $0.79 |
| | | | Yes | 48.39% | 52.41% | 55.1% | 72.35% | 60.71% | 3.57% | 0.61 hrs | $2.25 |
| | | QLoRA | No | 52.53% | 57.24% | 52.05% | 70.51% | 51.22% | 1.8% | 1.89 hrs | $6.93 |
| | | | Yes | 52.53% | 52.33% | 52.64% | 71.43% | 53.03% | 1.67% | 2.39 hrs | $8.78 |
| | Five-shot | None | No | 47.0% | 47.79% | 54.99% | 68.2% | 44.83% | 1.48% | 0.3 hrs | $1.08 |
| | | | Yes | 44.24% | 50.0% | 53.89% | 70.05% | 51.85% | 2.17% | 0.65 hrs | $2.37 |
| | | QLoRA | No | 41.47% | 44.35% | 47.83% | 57.6% | 27.59% | 2.75% | 2.05 hrs | $7.52 |
| | | | Yes | 46.08% | 46.94% | 49.72% | 65.44% | 42.11% | 2.54% | 2.6 hrs | $9.53 |
| Meta-Llama-3.1-70B-Instruct | Zero-shot | None | No | 68.2% | 70.61% | 69.43% | 85.71% | 80.7% | 1.43% | 0.21 hrs | $0.78 |
| | | | Yes | 64.98% | 66.7% | 64.1% | 82.95% | 68.83% | 0.79% | 1.12 hrs | $4.13 |
| | | QLoRA | No | 72.35% | 74.96% | 71.81% | 85.25% | 68.89% | 0.81% | 4.39 hrs | $16.11 |
| | | | Yes | 74.19% | 73.8% | 71.09% | 89.4% | 76.54% | 1.52% | 5.12 hrs | $18.78 |
| | Five-shot | None | No | 74.19% | 75.23% | 73.29% | 88.02% | 76.32% | 1.5% | 0.33 hrs | $1.20 |
| | | | Yes | 71.43% | 73.63% | 71.49% | 85.25% | 70.24% | 2.38% | 0.83 hrs | $3.06 |
| | | QLoRA | No | 62.67% | 72.77% | 63.41% | 74.19% | 54.39% | 3.03% | 4.57 hrs | $16.78 |
| | | | Yes | 66.36% | 74.12% | 65.34% | 77.88% | 58.04% | 0.0% | 5.38 hrs | $19.74 |
| | Zero-shot | None | No | 73.73% | 77.49% | 75.04% | 86.18% | 70.93% | 1.59% | 0.49 hrs | $1.78 |

| | | | | | | | | | | | |
|---|---|---|---|---|---|---|---|---|---|---|---|
| Mistral-Large-Instruct-2407 | | | Yes | 64.98% | 75.49% | 67.02% | 73.27% | 53.45% | 1.03% | 1.61 hrs | $5.91 |
| | Five-shot | None | No | 67.74% | 74.59% | 68.77% | 79.72% | 60.38% | 1.83% | 0.66 hrs | $2.41 |
| | | | Yes | 66.36% | 73.62% | 67.87% | 79.72% | 60.38% | 1.83% | 1.67 hrs | $6.13 |
| GPT-4o-2024-05-13 | Zero-shot | None | No | 82.95% | 81.1% | 81.71% | 91.24% | 80.52% | 0.74% | 0.05 hrs | $2.77 |
| | | | Yes | 75.58% | 77.61% | 74.48% | 86.18% | 70.0% | 3.23% | 0.11 hrs | $3.02 |
| | Five-shot | None | No | 78.8% | 77.08% | 76.38% | 89.4% | 76.54% | 0.76% | 0.1 hrs | $6.82 |
| | | | Yes | 72.35% | 75.37% | 72.2% | 82.95% | 64.95% | 2.56% | 0.1 hrs | $7.52 |

**Table S4:** Full unabridged table of statistically significant effects as well as intercepts across all four modeled performance metrics. We also show statistically significant estimated pairwise differences corresponding to statistically significant effects using Tukey's HSD.

| Response | Value | Variables and Pairwise Differences | Estimate |
|---|---|---|---|
| **Accuracy** | **Estimates** | (Intercept) | 57.3 |
| | | QLoRA | 15.6 |
| | | QLoRA:Five-shot | -17.7 |
| | **Estimated pairwise differences** | (QLoRA) – (No QLoRA) | 8.7 |
| | | (QLoRA, Zero-shot) – (No QLoRA, Zero-shot) | 16.6 |
| | | (QLoRA, Zero-shot) – (No QLoRA, Five-shot) | 14.8 |
| | | (QLoRA, Zero-shot) – (QLoRA, Five-shot) | 14.1 |
| **Precision** | **Estimates** | (Intercept) | 62.2 |
| | | QLoRA | 10.9 |
| | | QLoRA:Five-shot | -11.6 |
| | **Estimated pairwise differences** | (QLoRA) – (No QLoRA) | 6.7 |
| | | (QLoRA, Zero-shot) – (No QLoRA, Zero-shot) | 12.3 |
| | | (QLoRA, Zero-shot) – (No QLoRA, Five-shot) | 11.3 |
| | | (QLoRA, Zero-shot) – (QLoRA, Five-shot) | 10.2 |
| **Recall** | **Estimates** | (Intercept) | 62.4 |
| | | QLoRA | 8.3 |
| | | CoT | -6.2 |
| | | QLoRA:CoT | 6.8 |
| | | QLoRA:Five-shot | -11.9 |
| | **Estimated pairwise differences** | (No QLoRA) – (QLoRA) | 5.4 |
| | | (QLoRA, Zero-shot) – (No QLoRA, Zero-shot) | 11.7 |
| | | (QLoRA, Zero-shot) – (No QLoRA, Five-shot) | 9.7 |
| | | (QLoRA, Zero-shot) – (QLoRA, Five-shot) | 10.5 |
| | | (No QLoRA, No CoT) – (No QLoRA, CoT) | 4.2 |
| | | (QLoRA, No CoT) – (No QLoRA, CoT) | 4.4 |
| | | (QLoRA, No CoT) – (No QLoRA, CoT) | 6.5 |
| | | (QLoRA, CoT) – (No QLoRA, CoT) | 8.5 |

| | | | |
|---|---|---|---|
| **MES Scorable Accuracy** | **Estimates** | (Intercept) | 78.8 |
| | | QLoRA | 11.2 |
| | | QLoRA:Five-shot | -12.7 |
| | **Estimated pairwise differences** | (QLoRA) – (No QLoRA) | 6.8 |
| | | (QLoRA, Zero-shot) – (No QLoRA, Zero-shot) | 13.1 |
| | | (QLoRA, Zero-shot) – (No QLoRA, Five-shot) | 13.6 |
| | | (QLoRA, Zero-shot) – (QLoRA, Five-shot) | 13.1 |

**Table S5:** Three general prompt conversation templates are shown below. The annotation protocol provided to each model as system content, or first user content, is provided. Notice the many nuances in annotation for the Mayo endoscopic score. Additionally, templates for completion with and without chain-of-thought completion are presented.

| Template | Text |
|---|---|
| Annotation Protocol | ***MAYO ENDOSCOPIC SUBSCORE PROTOCOL***<br>**[PROTOCOL START]**<br><br>## Mayo Scorability Exclusion/Inclusion Criteria:<br>1. If a patient had a protocolectomy the report should NOT be annotated (a total abdominal colectomy is acceptable for annotation to enable scoring of the Hartmann's pouch). (Label as '-1'.)<br>2. If a procedure had no mucosa visualized due to a poor bowel preparation the report should NOT be annotated. (Label as '-1'.)<br>3. If a patient has a clear diagnosis of Crohn's disease the report should NOT be annotated. (Label as '-1'.)<br>- For example, Crohn's appears in the impressions section, or it appears in the indications section only and the findings to do not seem to question diagnosis. (Label as '-1'.)<br>4. If the patient carries a diagnosis of UC but has Crohn's disease like features (stricture, fistulae, aphthae), and impressions section does not clearly indicate a revised diagnosis from ulcerative colitis to Crohn's Disease, then the report CAN be annotated. (Provide Mayo score)<br>5. If the patient carries a diagnosis of 'IBD' then the report CAN be annotated so long as there is no mention of the patient having 'stricture(s)' and/or 'fistula(s)'. (Provide Mayo score)<br><br>A colonoscopy procedure report that cannot be annotated must be labeled as '-1'.<br><br>## Mayo Score:<br>If a report can be annotated, then you will score the report with a Mayo score. In particular, a Mayo score for a patient with ulcerative colitis will be given a Mayo score according to he most severely affected segment of the visualized colorectum according to the following descriptors:<br>1. Score a colonoscopy report with '0' for mentions of 'normal' (normal appearance), 'quiescent', 'scar' without other descriptors in the colorectum consistent with other classes.<br>2. Score a colonoscopy report with '1' for mentions of 'erythema', 'decreased vascular pattern', 'granularity', 'aphthous ulcer', 'aphthae', 'mild', no friability<br>3. Score a colonoscopy report with '2' for mentions of 'friability', marked or extensive 'erythema', 'loss of vascularity (absence)', 'erosions' (these can adjective mentions additionally distinguished with 'moderate').<br>4. Score a colonoscopy report with '3' for mentions of spontaneous 'bleeding', 'ulcers/ulcerated/ulceration', or 'severe' conditions.<br><br>## Mayo Score Edge Case:<br>Be mindful of the following edge cases:<br>1. In the case of poor bowel prep only score the report amongst the segments that are seen according to the previously mentioned scoring protocol.<br>2. If the the colonoscopy report indicates 'moderate-to-severe' but without any other descriptors of severity score the report with '3'.<br>3. If a clinician assignment of a Mayo score is discordant with other descriptors (e.g., report describes severity as mild but indicates presence of an ulcer), score the report with the most severe descriptor (e.g., 'severe colitis' with 'erosions' will be scored as '3', and 'mild' colitis with an 'ulcer' will be scored as '3').<br>4. For mentions of incomplete colonoscopy, sigmoidoscopy or proctoscopy (e.g., Hartmann's pouch) if the exam was intentionally halted due to distal disease severity, score as a 3. Otherwise, score only the segments that are seen.<br>6. IBD patients with...<br>- 'aphthous ulcers', score with a '1'.<br>- 'SCAD (erythematous mucosa), score with a '1'<br>- 'granular mucosa', score with a '3'.<br>- 'deep', 'shallow' or fissuring' ulcer,' score with a '3'.<br>- 'deep erosion', score with a '2'.<br>- 'scar/scarring', if described as associated with loss of vascularity, score with at least a '1'. Otherwise, the report cannot be annotated and should be labeled, '-1'.<br>- 'mucosal healing', ignore this descriptor and look for other descriptors to classify the severity.<br>- 'no active inflammation', ignore this descriptor and look for other descriptors to classify the severity.<br>7. If a patient does not a have a clear diagnosis of IBD (e.g., could be a screening colonoscopy or a diagnostic exam), so long as no exclusion criteria are met we can assign a Mayo score per the general rules (e.g., a clinical diagnosis |

| | |
|---|---|
| | of ulcerative colitis may be subsequently assigned)<br><br>## Additional Instruction<br>Make sure to think step-by-step in your annotation relating all decisions back to this annotation protocol and the contents of the colonoscopy procedure report. Again, be mindful of whether the patient can be given a Mayo score (i.e., they should have score "-1").<br>**[PROTOCOL END]** |
| Completion Only | ***IMPORTANT COMPLETION INSTRUCTION***<br>Provide the annotation of the COLONOSCOPY PROCEDURE REPORT included above strictly in JSON text as follows:<br><br>"{{<br>\"score\" : ...<br>}}"<br><br>where "score" can be '-1', '0', '1', '2' or '3' corresponding to the annotation protocol. Notice that the output is a JSON string and nothing else before or after. Follow this convention strictly and refrain from adding any other commentary before or after JSON output. |
| With Zero-shot-CoT | ***IMPORTANT COMPLETION INSTRUCTION***<br>Provide the annotation of the COLONOSCOPY PROCEDURE REPORT included above strictly in JSON text as follows:<br><br>"{{<br>\"score\" : ...,<br>\"reasoning\": ...<br>}}"<br><br>where "score" can be '-1', '0', '1', '2' or '3' corresponding to the annotation protocol and "reasoning" is a chain of thought explanation for the annotation that you provide. Notice that the output is a JSON string and nothing else before or after. Follow this convention strictly and refrain from adding any other commentary before or after JSON output. |
